# PRV-101 Coxsackievirus B vaccine elicits protective T follicular helper immunity while avoiding cytotoxic T-cell responses in humans: implications for type 1 diabetes prevention

**DOI:** 10.64898/2026.05.19.26352997

**Authors:** Federica Vecchio, Margot Petit, Orlando Burgos-Morales, Jutta E. Laiho, Mika Scheinin, Mikael Knip, Francisco León, Miguel Sanjuan, Heikki Hyöty, Sylvaine You, Roberto Mallone

**Affiliations:** Université Paris Cité, Institut Cochin, CNRS, INSERM, Paris, France; Faculty of Medicine and Health Technology, Tampere University, Tampere, Finland; Clinical Research Services Turku (CRST), LINK Medical, Turku, Finland; Instute of Biomedicine, University of Turku, Turku, Finland; Research Program for Clinical and Molecular Metabolism, Faculty of Medicine, University of Helsinki, Helsinki, Finland; Department of Pediatrics, Tampere University Hospital, Tampere, Finland; Turku Bioscience Centre, University of Turku and Åbo Akademi University, Turku, Finland; Provention Bio Inc., a Sanofi Company, Bridgewater, NJ, USA; Fimlab Laboratories, Tampere, Finland; Indiana Biosciences Research Institute, Indianapolis, IN, USA; Assistance Publique Hôpitaux de Paris, Service de Diabétologie et Immunologie Clinique, Cochin Hospital, Paris, France

## Abstract

PRV-101 is a multivalent formalin-inactivated Coxsackievirus B (CVB) vaccine developed to prevent CVB infections, which are associated with increased risk of islet autoimmunity. While PRV-101 induces robust neutralizing antibody responses, its T-cell immunogenicity is unknown. We analyzed peripheral blood mononuclear cells from 25 healthy adults receiving three high or low PRV-101 doses or placebo in a Phase I randomized, placebo-controlled trial. CVB-reactive CD8⁺ T-cell responses were assessed using HLA Class I multimers, and CD4⁺ and T follicular helper (Tfh) responses were measured by activation-induced marker assays following stimulation with a CVB peptide library. PRV-101 elicited minimal CVB-reactive CD8⁺ T-cell responses but robust CD4⁺ and Tfh responses, peaking at week 12 and persisting through week 32. Responses were observed in both seronegative and seropositive individuals, consistent with effective immune priming and boosting. Tfh frequencies correlated with neutralizing antibody titers. Female participants exhibited higher peak Tfh responses than males. We conclude that PRV-101 elicits a CVB-protective immune profile, dominated by Tfh responses supporting durable humoral immunity and devoid of potentially diabetogenic cytotoxic T-cell responses. This profile invites further investigations in vaccine trials for type 1 diabetes prevention.

**Article Highlights:** PRV-101 induces robust neutralizing antibody responses, but its cellular immunogenicity remains undefined. Given the role of CD4⁺ T follicular helper (Tfh) cells in sustaining humoral immunity, we asked whether PRV-101 induces protective T-cell responses. We found that PRV-101 elicited minimal CD8⁺ T-cell responses but robust Tfh responses in both CVB-seronegative and seropositive individuals, which correlated with neutralizing antibody titers. These findings indicate a coordinated humoral and Tfh response deviated away from potentially diabetogenic cytotoxic CD8⁺ T-cell activation, supporting further evaluation of PRV-101 for type 1 diabetes prevention.

---

Type 1 diabetes (T1D) results from the autoimmune destruction of pancreatic beta cells, leading to lifelong insulin therapy for survival. Currently, no cure or preventive treatments exist. While genetic factors, particularly HLA Class II haplotypes, confer disease susceptibility, the rising global incidence of T1D - even in individuals with neutral or protective HLA alleles - strongly implicates environmental triggers (1). Among these, enteroviruses, particularly viruses belonging to Coxsackievirus B (CVB), have emerged as prime candidates based on histopathological, epidemiological, and mechanistic evidence (2,3).

CVB comprises six serotypes (CVB1-6) within the *Enterovirus* genus. These non-enveloped RNA viruses cause diseases ranging from mild gastroenteritis to severe myocarditis and meningitis, and may initiate the autoimmune process that precedes T1D clinical onset (4). Indeed, multiple lines of evidence link CVB to T1D pathogenesis. Enteroviral proteins and RNA have been detected in pancreatic islets and the gut of individuals with T1D (5,6), often colocalizing, in the pancreas, with HLA Class I hyperexpression, hallmark of the disease (7,8). Prospective studies demonstrate that CVB infections, and CVB infections with prolonged (persistent) shedding of the virus, in genetically at-risk children correlate with islet auto-antibody seroconversion, marking autoimmunity initiation (9–11). These persistent infections may reflect inefficient antiviral responses, as observed in children who seroconvert for IAA within the first years of life (12), potentially facilitating pancreatic spreading and subsequent autoimmunity (11,13). Moreover, antiviral treatment in new-onset type 1 diabetic children and adolescents has recently shown positive results in maintaining residual beta-cell function (14), suggesting a need to extend these antiviral strategies to T1D prevention trials.

PRV-101 is a multivalent formalin-inactivated CVB vaccine targeting CVB serotypes 1-5. A recent Phase I trial in healthy adults demonstrated that PRV-101 is well tolerated and induces robust neutralizing antibody (Ab) responses across all serotypes (15), even after a prolonged period of time (16). However, cellular immune responses, particularly T-cell-mediated immunity, are critical determinants of vaccine efficacy and safety. Natural CVB infection induces differential T-cell responses (17), characterized by robust CD4⁺ T helper activation but limited CD8⁺ T-cell responses, which are marked by developmental arrest and exhaustion features (13,18). Recently, we have further confirmed this divergent T-cell immunity pattern in enterocytes and antigen-presenting cells, where CVB3 infection leads to HLA Class I downregulation impairing CD8^+^ T-cell responses, while HLA Class II upregulation supports efficient polyfunctional CD4^+^ T-cell differentiation (18). Whether PRV-101 vaccination recapitulates or diverges from this T-cell pattern is essential for predicting protective immunity.

T follicular helper (Tfh) cells are detectable in the blood and represent specialized CD4⁺ T cells that express CXCR5, which directs their migration to B-cell follicles (19). Within germinal centers, Tfh cells provide critical B-cell help through CD40L-CD40 interactions and the secretion of IL-21, driving Ab class switching, somatic hypermutation, and the generation of long-lived plasma cells (20). Thus, Tfh response magnitude and quality are intimately linked to vaccine-induced Ab durability and breadth. In parallel with Tfh-mediated support of humoral immunity, cytotoxic CD8^+^ T cells represent another critical component of antiviral defense by recognizing viral peptides on HLA Class I molecules and eliminating infected cells. While crucial for clearing intracellular pathogens, excessive CD8⁺ T-cell activation could theoretically contribute to beta-cell immunopathology through direct cytotoxicity, bystander damage, or epitope spreading (3).

Such immune mechanisms do not operate uniformly across individuals. Sex differences in vaccine responses are well documented, with females generally mounting more robust Ab and cellular responses than males (21–23). Understanding sex-specific vaccine responses is crucial for optimizing vaccination regimens. Together, these considerations underscore the need for a comprehensive evaluation of vaccine-induced cellular immunity.

Here, we conducted a comprehensive immunological analysis of T-cell responses elicited by PRV-101 in healthy adults participating in a Phase I trial (15). Our objectives were to: (1) characterize the magnitude and kinetics of vaccine-induced CD4⁺ and CD8⁺ T-cell responses; (2) assess whether PRV-101 preferentially activates Tfh cells supporting humoral immunity; (3) identify sex-specific differences in immunogenicity; and (4) compare the T-cell profiles induced by vaccination vs. those induced by natural CVB infection.

## RESEARCH DESIGN AND METHODS

### Study design and participants

This immunological ancillary study analyzed specimens from the PRV-101 Phase I clinical trial (PROVENT trial, NCT04690426), a double-blind, randomized, placebo-controlled study evaluating multiple-dose-escalation PRV-101 safety and immunogenicity in healthy adults (15). The parent trial enrolled 32 participants aged 18-45 years at Clinical Research Services, Turku, Finland. Main inclusion criteria were age 18-45 years and good general health. Exclusion criteria included: severe allergic reactions; immunodeficiency or immunosuppressive therapy; autoimmune disease; pregnancy or breastfeeding; and participation in other investigational studies.

PRV-101 composition, manufacturing, and administration procedures have been previously described (15). Briefly, participants received three intramuscular injections of either high-dose (500 μl) or low-dose (100 μl) PRV-101, or placebo at weeks 0, 4, and 8. Participants were randomized 3:1 (PRV-101:placebo) and all study personnel, including those performing immunological analyses in this study, remained blinded to treatment allocation until primary analyses were completed.

The study was approved by the Ethics Committee of the Hospital District of Southwest Finland and conducted according to Good Clinical Practice guidelines and Declaration of Helsinki principles. All participants provided written informed consent. Immunological analyses were performed on samples from 25 of the 32 participants, selected based on HLA-A*02:01^+^ or HLA-A*03:01^+^ status and sample availability: high-dose PRV-101 (n=10), low-dose PRV-101 (n=7), and placebo (n=8).

### Blood samples

Peripheral venous blood (40-50 ml) was collected in sodium heparin tubes at weeks 0 (baseline), 4, 8, 12, and 32. Peripheral blood mononuclear cells (PBMCs) were isolated using BD Vacutainer® CPT™ (Mononuclear Cell Preparation) sodium citrate tubes according to manufacturer’s instructions, cryopreserved in FBS with 10% DMSO, and stored in liquid nitrogen according to standardized protocols (24).

### CVB serostatus and HLA typing

CVB serostatus was determined by neutralizing plaque reduction assay measuring neutralizing Abs against CVB1-5, as previously described (15). Titers ≥1:8 were considered positive. Participants with detectable Abs to at least one serotype at baseline were classified as CVB-seropositive. Genotyping data for HLA-A*02:01 (HLA-A2 from hereon; n=12 participants), HLA-A*03:01 (HLA-A3 from hereon; n=12 participants) and HLA-A2/A3 (n=1) from the parent trial were used for allocation to HLA Class I multimer studies.

### HLA Class I multimer assays

CVB peptides were synthesized to >85% purity (Synpeptide), following selection based on our recent immunopeptidomics and T-cell studies (13), prioritizing sequences with high conservation across CVB serotypes. Structural epitopes were derived from VP1-4 capsid proteins (P1 region); non-structural epitopes were derived from replication proteins (P2-P3 regions).

HLA-A2-restricted peptides included CVB5_169-177_ (YLGRAGYTV, structural) and CVB1_1671-1679_ (RMLMYNFPT, non-structural). HLA-A3-restricted peptides included CVB5_760-768_ (SMFYDGWAK, structural) and CVB1_1356-1364_ (KINMPMSVK, non-structural). Sequence conservation across serotypes is depicted in Supplementary Fig. 1A.

HLA Class I multimers were generated as previously described (13). Biotinylated HLA-A*02:01 or HLA-A*03:01 heavy chain and beta2-microglobulin were folded with peptides and multimerized using fluorochrome-conjugated streptavidin (PE, APC, BV786; BD Biosciences).

Frozen PBMCs were thawed in pre-warmed AIM-V medium (ThermoFisher), and CD8⁺ T cells were enriched by negative magnetic selection (EasySep, Stemcell Technologies). Enriched cells (2-5×10^6^) were stained with combinatorial double-coded multimer panels for 20 min at RT in 20 μl of PBS-dasatinib for 10^7^ cells followed, without washing, by staining at 4°C for 20 min with monoclonal (m)Abs CD3-APC-H7 (RRID:AB_1645475), CD8-PE-Cy7 (RRID:AB_396852), and Live/Dead Aqua (ThermoFisher L34957). At least 1-2×10⁶ CD8⁺ T cells per sample were acquired on a Cytek Aurora spectral flow cytometer. Data were analyzed using FlowJo v10.8. Frequencies were calculated as multimer⁺ cells out of total CD8⁺ T cells.

### Design of CVB P1 structural protein peptide library

To comprehensively assess CD4⁺ T-cell responses to CVB structural antigens in vaccinated participants, we designed an overlapping peptide library spanning the entire CVB P1 region (capsid polyprotein). The CVB3 P1 polyprotein sequence (848 amino acids, aa) served as the reference, generating 168 overlapping 15-mer peptides with 10-aa overlaps. The peptide library was optimized using a multi-serotype alignment strategy: the P1 sequences from the 5 PRV-101 CVB strains (CVB1 to 5) were compared at each position, and the most conserved aa was selected. When all five strains showed sequence variation, the CVB3 aa was retained. When the CVB3 residue differed from the other four strains but two strains shared an alternative residue, the CVB1 aa was selected, prioritizing the two most prevalent serotypes in the general population (CVB1 and CVB3). This strategy ensured representation of the most conserved and clinically relevant sequences across naturally circulating CVB strains.

#### Activation-induced marker (AIM) assays

Frozen PBMCs (8×10⁶) were thawed and cultured in 48-well plates with anti-CD40 Ab (RRID:AB_10839704; 1 µg/ml) to prevent CD40L downregulation (25). Cells were stimulated with the pooled CVB P1 overlapping peptide library at a final concentration of 1 µM per peptide for 24 h at 37°C, 5% CO₂, with non-stimulated wells serving as negative controls. Cells were then stained with Live/Dead Green viability marker (ThermoFisher L23101) and the following fluorochrome-conjugated mAbs: CD3-BUV496 (RRID:AB_2870222), CD8-BUV563 (RRID:AB_2870199), CD4-BUV805 (RRID:AB_2870176), CD25-BV711 (RRID:AB_2738037), CD69-PE/Dazzle594 (RRID:AB_2564277), HLA-DR-BUV661 (RRID:AB_2870252), OX40/CD134-PE/Cyanine5 (RRID:AB_2894615), CXCR5/CD185-AlexaFluor647 (RRID:AB_2737606), and CD40L/CD154-PE (RRID:AB_2751142). Cells were incubated with mAbs for 20 min at 4°C in the dark, then fixed and acquired on a Cytek Aurora spectral flow cytometer. At least 1×10⁶ CD4⁺ T cells were acquired per condition and analyzed using FlowJo v10.8. Activation-induced marker (AIM)⁺CD4⁺ T cells were identified using Boolean gating. For each sample, AIM combinations (CD154⁺CD25⁺CD69⁺, CD154⁺CD25⁺HLA-DR⁺, or CD154⁺CD25⁺OX40⁺) present in peptide-stimulated samples were defined as antigen-reactive and merged using OR gating (25). CXCR5⁺ Tfh cells were identified within the AIM⁺CD4⁺ population. Frequencies were calculated as AIM^+^ cells out of total CD4⁺ T cells.

### Statistical analysis

Sample size was dictated by the parent Phase I trial design. Statistical analyses were performed using GraphPad Prism v9.4. Individual data points are shown in each graph. Comparisons between two groups used the Mann-Whitney U test (unpaired) and multiple group comparisons used the Friedman test with Dunn’s correction. Correlations were assessed using Spearman’s rank correlation. Two-tailed p values <0.05 were considered statistically significant.

## RESULTS

### Study participants and baseline characteristics

Twenty-five healthy adults (median age 26 years, range 19-44) were included in this immunological sub-study (Table 1). Participants received three intramuscular PRV-101 doses (high or low) or placebo at weeks 0, 4 and 8, with follow-up through week 32 (Fig. 1A). Overall, seven participants (28%) were CVB-seronegative at baseline, with similar distributions across treatment arms: high-dose (37.5%), low-dose (14.3%) and placebo (30.0%) (Fig. 1B).

**Fig. 1.**
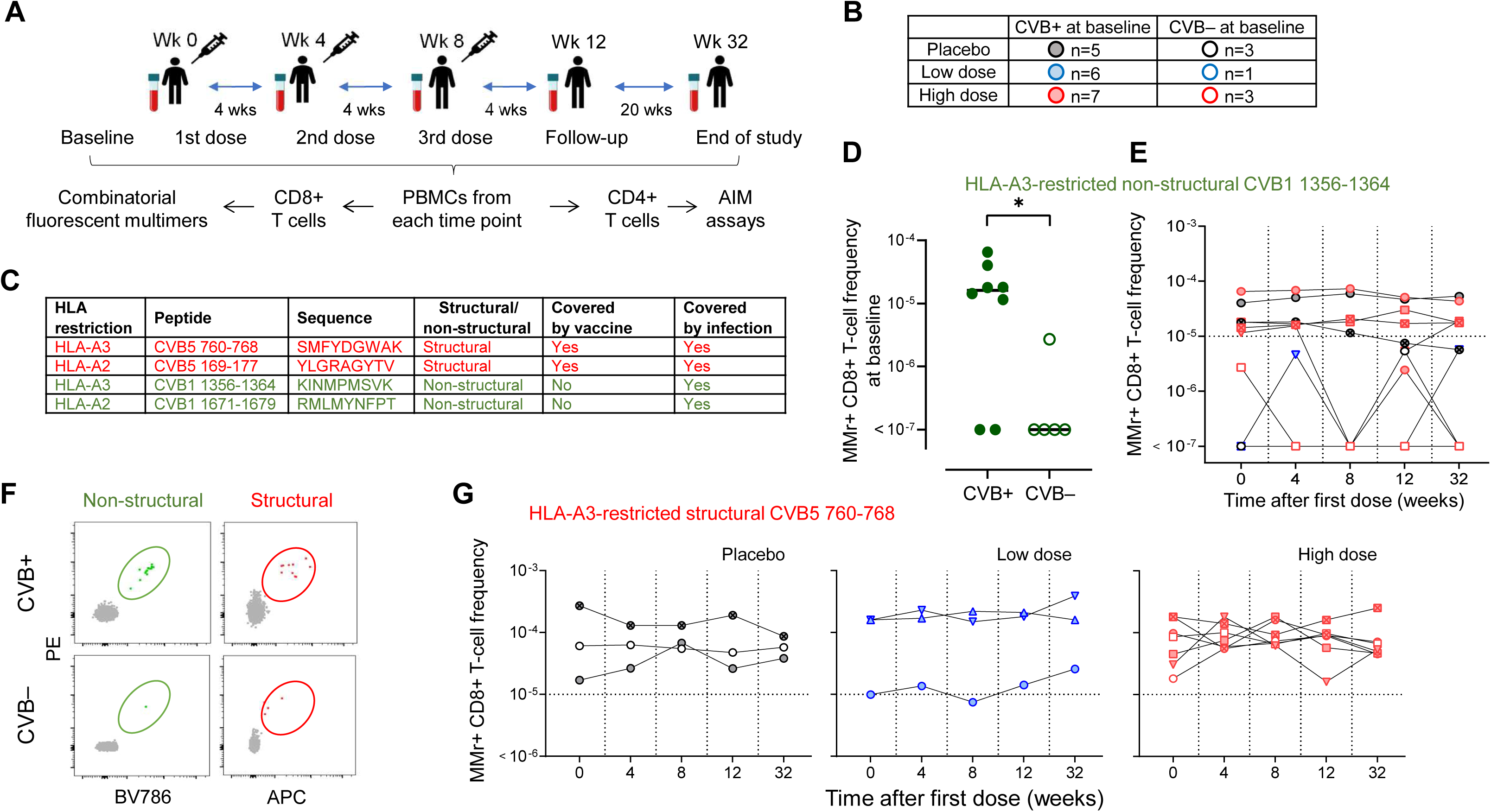
Study design and CD8⁺ T-cell responses to CVB epitopes. (**A**) Study outline showing vaccination schedule (three doses at weeks 0, 4, and 8), blood sampling time points (weeks 0, 4, 8, 12, and 32) and experimental workflow: PBMCs analyzed by combinatorial fluorescence multimer assays for CD8⁺ T cells and activation-induced marker (AIM) assays for CD4⁺ T cells. (**B**) Treatment allocation: high-dose PRV-101 (n=10; 7 CVB-seropositive, 3 CVB-seronegative), low-dose PRV-101 (n=7; 6 CVB-seropositive, 1 CVB-seronegative), and placebo (n=8; 5 CVB-seropositive, 3 CVB-seronegative). (**C**) CVB peptides used in multimer assays: HLA-A3-restricted and HLA-A2-restricted structural (red) and non-structural (green) epitopes. Non-structural peptide-reactive T-cell responses serve as biomarkers for natural infection; structural peptides are those covered by PRV-101 vaccine. (**D**) Frequencies of CD8⁺ T cells reactive to the HLA-A3-restricted non-structural CVB1_1356-1364_ epitope in CVB-seropositive (filled symbols) vs. CVB-seronegative (empty symbols) participants at baseline. (**E**) Frequencies of CD8⁺ T cells reactive to the same epitope across all timepoints and treatment arms. (**F**) Representative flow cytometry plots of multimer staining for HLA-A3-restricted non-structural (green) and structural (red) epitopes detected in CVB-seropositive and seronegative participants at baseline. (**G**) Frequencies of CD8⁺ T cells reactive to the HLA-A3-restricted structural CVB5_760-768_ epitope across placebo, low-dose, and high-dose treatment arms at all timepoints.

**Table 1.**
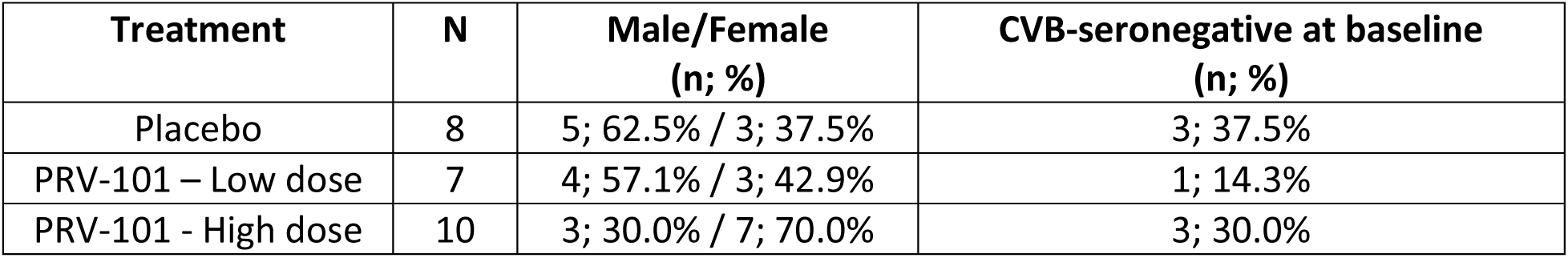
Baseline characteristics of PRV-101 vaccine trial participants included in this immunological analysis, stratified by treatment arm. CVB-seronegative status was assessed at baseline visit (week 0), prior to vaccination. A seropositive status was assigned in the presence of neutralizing Abs against ≥1 CVB serotype. No significant differences between groups (p>0.3 for all comparisons by Kruskal-Wallis or Fisher’s exact test).

### CD8⁺ T-cell responses to non-structural epitopes distinguish CVB-seropositive from CVB-seronegative individuals, but not vaccination status

To validate CVB serostatus and assess pre-existing cellular immunity, we examined baseline CD8⁺ T-cell responses using HLA Class I multimers loaded with CVB epitopes. We included both structural epitopes (derived from VP1-4 capsid proteins present in inactivated PRV-101) and non-structural epitopes (derived from replication proteins expressed only during active viral replication, serving as natural infection biomarkers) (Fig. 1C). For HLA-A2⁺ participants (n=12), we used structural epitope CVB5_169-177_ and non-structural epitope CVB1_1671-1679_. For HLA-A3⁺ participants (n=12), we used structural epitope CVB5_760-768_ and non-structural epitope CVB1_1356-1364_. All epitopes showed >95% sequence conservation across CVB serotypes (Supplementary Fig. 1A). At baseline (Fig. 1D), CVB non-structural peptide-reactive CD8⁺ T cells were readily detectable in HLA-A3^+^ CVB-seropositive individuals (median frequencies 1.61×10^-5^ CD8⁺ T cells for HLA-A3-restricted CVB1_1356-1364_), but significantly lower or undetectable in CVB-seronegative individuals (median 1×10^-7^, P= 0.0326), confirming that cellular immunity to non-structural proteins reflects prior CVB exposure. These frequencies remained stable throughout follow-up (Fig. 1E), indicating absence of novel CVB infections during the trial.

Irrespective of treatment allocation, CD8^+^ T-cell responses against structural peptides (included in the vaccine) were also stable across time points in all three treatment arms (placebo, low dose and high dose) (Fig. 1F-G). Similar results were obtained for HLA-A2^+^ participants (Supplementary Fig. 1B). These results demonstrate that, unlike live viral infections, PRV-101 does not elicit significant cytotoxic CD8⁺ T-cell responses to structural vaccine antigens.

### PRV-101 elicits robust CD4⁺ Tfh responses

Given this limited CD8⁺ T-cell activation, we hypothesized that PRV-101 might preferentially induce CD4⁺ T helper responses, particularly Tfh cells, crucial for B-cell responses and Ab production. We employed an activation-induced marker (AIM) assay (25) identifying antigen-reactive CD4⁺ T cells based on co-expression of AIMs following peptide stimulation. PBMCs collected at each time point were stimulated for 24 h with overlapping peptide pools spanning CVB structural proteins (included in the vaccine). Following stimulation, cells were stained for CD3, CD4, CXCR5 (Tfh marker) and AIMs, including CD154 (CD40L), CD25, CD69, HLA-DR and OX40 (CD134). We employed Boolean gating to identify AIM⁺CD4⁺ T cells based on combinations such as CD154⁺CD25⁺OX40⁺, CD154⁺CD25⁺HLA-DR⁺ and CD154⁺CD25⁺CD69⁺, maximizing sensitivity and specificity for rare antigen-reactive T cells (Fig. 2A).

**Fig. 2.**
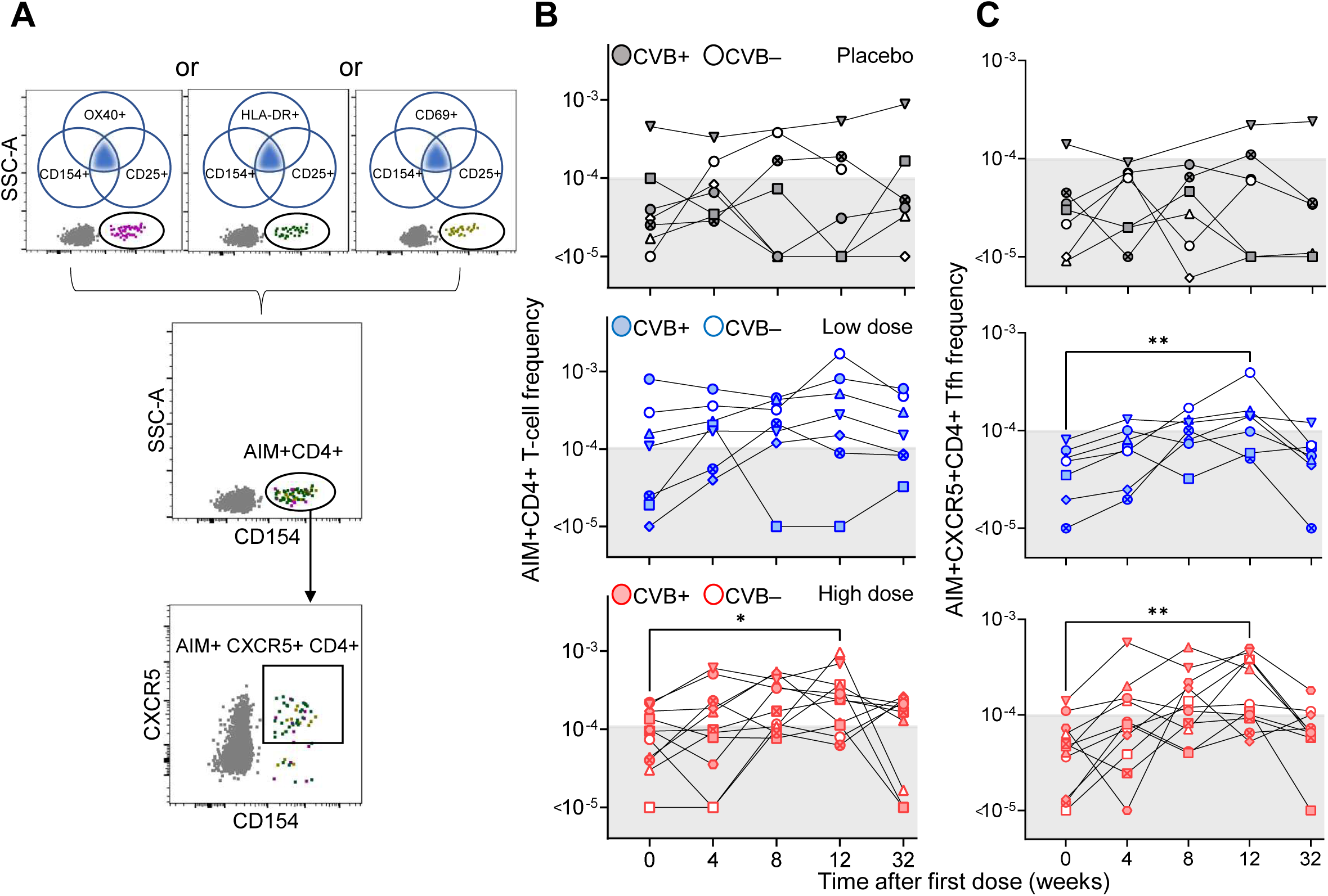
PRV-101 elicits robust CD4⁺ Tfh responses. (**A**) Boolean gating strategy for identifying AIM⁺CD4⁺ T cells using combinations of CD154 with CD25 and OX40, HLA-DR or CD69. Representative staining of activated (AIM^+^) CD4^+^ T cells and Tfh cells (AIM^+^CXCR5^+^CD4^+^). (**B-C**) Longitudinal frequencies of CVB structural peptide-reactive AIM^+^CD4^+^ T cells (B) and AIM^+^CXCR5^+^CD4^+^ Tfh cells (C) across placebo (grey, top panel), low-dose (blue, middle panel), and high-dose (red, bottom panel) treatment arms. CVB-seropositive (filled symbols) and CVB-seronegative participants (open symbols) are shown separately. Each symbol represents one participant. *p<0.05, **p<0.01 by Tukey’s multiple comparison test applied to mixed effects model.

At baseline, CVB structural peptide-reactive AIM⁺CD4⁺ T cells were detectable in both CVB-seropositive and seronegative participants (Fig. 2B). Following PRV-101 vaccination, we observed marked and progressive increases in CVB structural peptide-reactive AIM⁺CD4⁺ T cells in the high-dose arm (Fig. 2B). A similar, although not significant, increase was also observed in the low-dose treatment arm, but not in the placebo arm. This increase was evident by week 8 (i.e., after the second dose) and continued to rise through week 12 (i.e., after the third dose), decreasing yet remaining above baseline values at week 32 in most participants. In the high-dose arm, median frequencies increased from baseline (9.4×10^-5^ CD4⁺ T cells) to peak at week 12 (2.4×10^-4^, p=0.0267) and persisting at week 32 (1.7×10^-4^). The low-dose arm showed similar patterns, with a non-significant yet notable increase by week 12 (2.8×10^-4^ vs. 1.1×10^-4^ at baseline) although not homogeneously persisting at week 32 (1.5×10^-4^). In contrast, no significant changes were noted in the placebo arm.

To determine whether vaccine-induced CD4⁺ T cells acquired Tfh characteristics, we analyzed CXCR5 expression within the AIM⁺CD4⁺ population. Following PRV-101 vaccination, we observed a more pronounced increase in CXCR5⁺ T-cell frequencies within the AIM⁺CD4⁺ population (Fig. 2C). In the high-dose arm, median AIM⁺CXCR5⁺CD4⁺ Tfh cell frequencies increased significantly by week 12 (1.0×10^-4^ vs 4.7×10^-5^ at baseline, p=0.0015). The low-dose arm displayed similar trends (1.4×10^-4^ vs 4.8×10^-5^ at baseline, p= 0.0078), while the placebo arm showed no significant change. In both the high-and low-dose arms, AIM⁺CXCR5⁺CD4⁺ Tfh cell frequencies remained elevated at week 32 (5.5×10^-5^ and 6.7×10^-5^, respectively), barring one participant in each group.

These results demonstrate that PRV-101 induces robust, dose-dependent CD4⁺ Tfh responses to vaccine-targeted structural antigens, with responses detectable after the first dose and peaking after the third dose.

### Vaccine-induced Tfh responses correlate with neutralizing antibody titers in both CVB-seropositive and seronegative individuals

To assess protective correlates of PRV-101-induced Tfh responses, we examined relationships between AIM⁺CXCR5⁺CD4⁺ T-cell frequencies and neutralizing Ab titers. One representative example is shown in Fig. 3A. Tfh responses induced by PRV-101 paralleled neutralizing Ab responses, with titers increasing progressively through week 12 and remaining elevated at week 32. This correlation was further analyzed by stratifying for CVB serostatus, identified as presence or absence of neutralizing CVB Abs at baseline. In CVB-seronegative participants (Fig. 3B), we observed that the increase in AIM⁺CXCR5⁺CD4⁺ Tfh frequencies paralleled that of neutralizing Ab titers. Although weak, this correlation was statistically significant. In CVB-seropositive participants (Fig. 3C), this correlation was also statistically significant although weaker, likely reflecting the confounding effect of pre-existing neutralizing Abs prior to vaccination. Accordingly, Ab titers were higher at both baseline and follow-up.

**Fig. 3.**
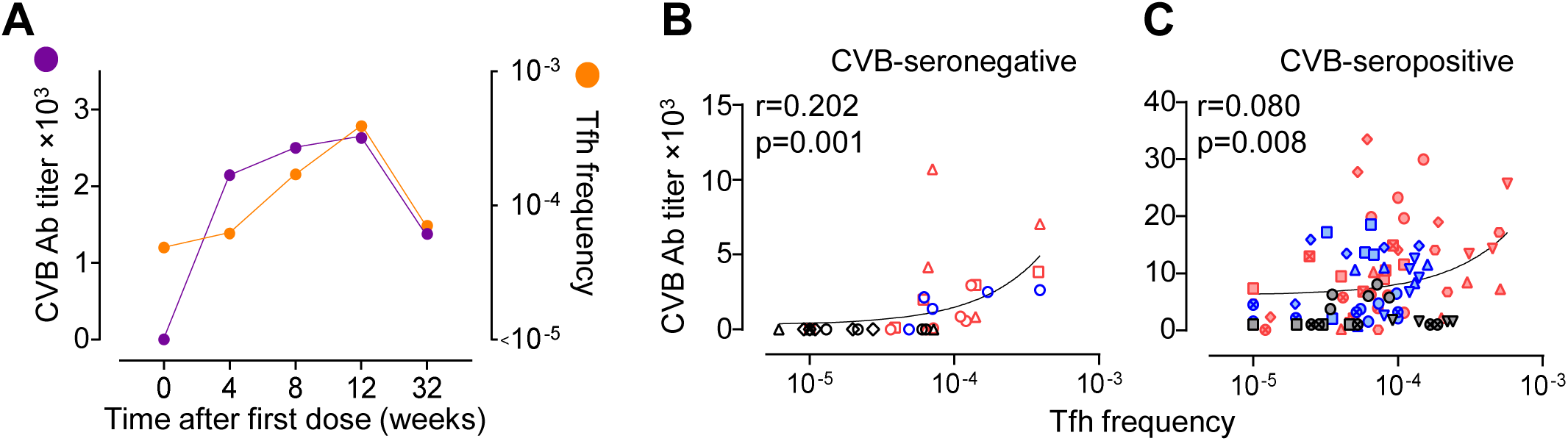
Relationship between vaccine-induced Tfh and neutralizing Ab responses against CVB. (**A**) Representative plot showing increase in CVB Ab titers (sum of Ab titers for all 5 PRV-101 serotypes) and AIM^+^ Tfh frequencies out of total CD4^+^ T cells for one low-dose CVB-seronegative PRV-101 recipient over time. (**B-C**) Scatter plots with linear regression showing the relationship between AIM⁺CXCR5⁺CD4⁺ Tfh cell frequency (x-axis) and total neutralizing CVB antibody titers summed across all 5 serotypes (y-axis) in CVB-seronegative (B) and CVB-seropositive (C) participants. Spearman correlation coefficients and p values are shown for each graph.

This Tfh-Ab relationship demonstrates that vaccine-induced Tfh cells are functionally coupled to neutralizing Ab production, supporting the notion that Tfh activation is a key vaccine-induced protection mechanism. Moreover, the PRV-101 vaccine can both prime naïve immune responses in CVB-seronegative individuals and boost pre-existing memory responses in CVB-seropositive individuals.

### Female participants exhibit significantly higher Tfh responses

To determine whether sex influences PRV-101 immunogenicity, we stratified analyses by participant sex. The cohort included 14 females and 11 males evenly distributed across treatment arms, with comparable baseline characteristics including age and CVB serostatus (Supplementary Table 1).

At baseline, CVB structural peptide-reactive AIM⁺CD4⁺ T-cell frequencies were similar between females and males in all treatment arms. Following PRV-101 vaccination, females exhibited overall significantly higher increase in CD4⁺ T-cell frequencies compared to males (Fig. 4A). In the high-dose PRV-101 arm, females showed significantly increased CVB-reactive AIM⁺CD4⁺ T-cell frequencies at weeks 12 compared to males. Similar although not statistically significant trends were observed in the low-dose arm, but not in the placebo arm.

**Fig. 4.**
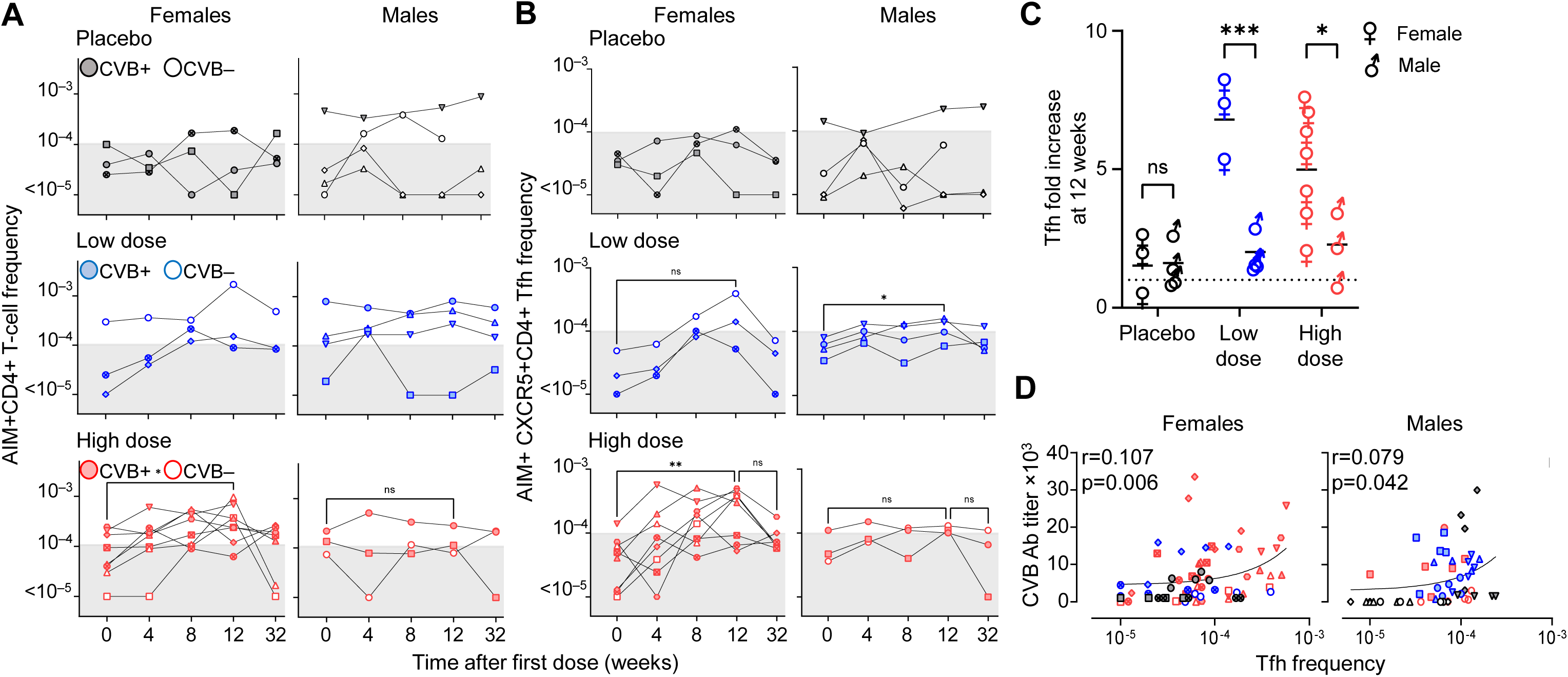
Vaccine-induced Tfh and neutralizing antibody responses against CVB stratified by sex. (**A-B**) Longitudinal frequencies of CVB structural peptide-reactive AIM⁺CD4⁺ T cells (A) and AIM^+^CXCR5^+^CD4^+^ Tfh cells (B) across weeks 0, 4, 8, 12, and 32, stratified by sex (left and right columns) and treatment arm (top, middle and bottom row). Each symbol represents one participant, with filled and open symbols indicating CVB-seropositive and CVB-seronegative status at baseline, respectively. *p<0.05, **p<0.01, ns = not significant by Tukey’s multiple comparison test applied to mixed effects model. (**C**) Tfh fold increase at 12 weeks across placebo, low-dose, and high-dose treatment arms in female vs. male participants. ***p=0.0004 and *p=0.015 by Dunnett’s multiple comparison test. (**D**) Scatter plots with linear regression showing relationship between AIM⁺CXCR5⁺CD4⁺ Tfh cell frequency (x-axis) and total neutralizing CVB Ab titers (y-axis) stratified by sex. Spearman correlation coefficients and p values are shown for each graph.

Sex differences were even more pronounced for Tfh responses (Fig. 4B). Overall, females exhibited higher increase in AIM⁺CXCR5⁺CD4⁺ T-cell frequencies at week 12 relative to their baseline as compared to males, with significant female to male differences both in the low-and in the high-dose arm (Fig. 4C). This pattern suggests that females mount more robust Tfh responses to PRV-101 vaccination. Accordingly, the correlation with neutralizing CVB Ab titers was more pronounced in females than in males (Fig. 4D).

These findings reveal sex-specific differences in PRV-101-induced Tfh responses that may guide vaccine dosing strategies.

## DISCUSSION

PRV-101 vaccination induced robust CVB-specific CD4⁺ Tfh responses associated with neutralizing CVB Ab production, providing mechanistic insights on the induction of protective immunity. This preferential Tfh response aligns with fundamental vaccine immunology principles. Unlike live-attenuated vaccines, inactivated formulations primarily engage indirect antigen presentation through antigen uptake, favoring HLA class II-restricted CD4⁺ T-cell activation (20). The intramuscular route of administration enhances antigen drainage to lymph nodes and efficient uptake by professional antigen-presenting cells in secondary lymphoid organs, preferentially inducing CD4⁺ T-cell responses (26,27). Our observation that vaccine-induced Tfh responses correlate with raise in neutralizing Ab titers suggests that PRV-101 establishes long-lived plasma cells and memory B cells, the cellular basis for durable humoral immunity (16). This may be further favored by the prime and boost vaccination scheme (three doses) and vaccine formulation, which together drive sustained germinal center reactions and affinity maturation.

PRV-101 did not induce CD8⁺ T-cell responses to structural epitopes. This finding has several important implications. First, it indicates that the engagement of HLA class I antigen presentation pathways is minimal. While cross-presentation – by which endocytosed antigens are presented on HLA class I molecules - can occur, it typically requires specific conditions lacking in the current PRV-101 formulation, such as adjuvants promoting dendritic-cell activation, or targeted delivery to cross-presenting dendritic cell subsets. Our recent proteomics and immunopeptidomics analysis of CVB infection in enterocytes (18) show that even natural CVB infection elicits limited CD8⁺ T-cell responses due to immune escape mechanisms, notably HLA class I downregulation, while HLA class II upregulation supports robust polyfunctional CD4⁺ T cell responses, including Tfh subsets. Second, lack of induction of CD8⁺ T-cell responses may represent a safety advantage in future T1D prevention trials. Natural CVB infection induces CD8⁺ T-cell responses characterized by developmental arrest in naïve and early memory states with exhaustion features (13,18). These impaired responses may paradoxically promote chronic inflammation through incomplete viral clearance. Moreover, the CD8⁺ T-cell response induced, although limited, may retain cytotoxic potential and contribute to beta-cell destruction through different mechanisms (3). These include direct killing of CVB-infected beta cells, although we showed that this is marginal due to viral escape mechanisms (13); bystander damage to uninfected beta cells through local inflammation; epitope spreading, wherein beta-cell antigens released during viral cytolysis prime autoreactive CD8⁺ T cells; and molecular mimicry between viral and self-epitopes. By avoiding CD8⁺ T-cell activation, PRV-101 may reduce this risk while maintaining protective Ab responses that neutralize CVB entry upstream of viral spreading and beta-cell infection. This concept is supported by observations that Ab-mediated protection is sufficient to prevent enterovirus-induced diabetes in animal models (28,29), and that neutralizing Abs correlate with protection against viral diseases in humans (30).

On the other hand, the absence of CD8⁺ T-cell responses raise questions about PRV-101’s ability to clear breakthrough infections. Mitigating this concern, PRV-101 induces robust Tfh and neutralizing Ab responses that should prevent infections *ab initio* and be boosted more rapidly upon natural exposure. Of further note, chronic low-grade infections rather than acute infections have been linked to development of islet auto-Abs (11). By providing high neutralizing Ab titers, PRV-101 could prevent the initial acute infection and thus also the formation of reservoirs nurturing viral persistence.

The PRV-101 vaccine demonstrates advantages over other enterovirus interventions. Unlike antiviral approaches that would need to be administered after infection (14), PRV-101 provides preventive protection. In this context, the choice between inactivated and live-attenuated vaccine platforms involves careful consideration of immunological and safety trade-offs. The extensive literature on oral poliovirus vaccines (OPV) versus inactivated poliovirus vaccines (IPV) provides instructive parallels. Live-attenuated vaccines such as OPV offer distinct advantages including robust mucosal immunity, ease of administration, and potential for herd immunity through secondary transmission (31). However, attenuated vaccine strains can undergo genetic reversion, particularly in under-immunized populations, potentially leading to vaccine-derived virulent variants (32). While significant progress has been made in engineering genetically stabilized live-attenuated vaccines to reduce reversion risk (33), this challenge remains an inherent consideration for live vaccines. Inactivated vaccines such as PRV-101 eliminate reversion risk entirely by using non-replicating viral antigens, though typically with reduced mucosal immunity compared with live-attenuated counterparts. Considering the ultimate objective of testing CVB vaccination for T1D prevention, the PRV-101 inactivated formulation prioritizes safety, given the target population of healthy at-risk children.

Sex-specific differences in vaccination immune correlates represents another interesting finding, with women mounting significantly higher Tfh responses than men. These sex differences in vaccine immunogenicity are well documented, with females generally mounting more robust Ab responses to various vaccines, e.g. influenza, hepatitis B, rabies and yellow fever (21–23). Estrogens enhance both B-cell and T-cell responses through multiple mechanisms: upregulation of B-cell survival factors, CD40L expression on CD4⁺ T cells promoting T/B-cell interactions, and IL-21 production (34–38). X-chromosome dosage effects are linked to incomplete X inactivation in females, leading to biallelic expression of immune genes such as TLR7, CD40L and CXCR3, which could enhance Tfh differentiation and function. Sex differences in vaccine immunogenicity should inform trial design and analysis plans, including prospective investigation of sex-stratified dosing strategies.

Altogether, these observations position PRV-101 vaccination as a safe option for testing its potential for T1D prevention. The rationale for such trials is strengthened by the observation that at-risk children developing islet auto-Abs during the first years of life lack neutralizing CVB Abs (12). The demonstration that PRV-101 successfully primes *de novo* Tfh and Ab responses in CVB-seronegative individuals while also boosting pre-existing immunity in CVB-seropositive participants indicates broad applicability across populations with varying prior CVB exposure. The vaccine may also have implications beyond T1D prevention. Besides mild yet significant clinical manifestations such as gastroenteritis and flu-like syndromes, CVB infection is a major etiologic factor of less frequent yet potentially life-threatening conditions in childhood, such as myocarditis and meningitis (15,39,40). The robust Ab responses induced by PRV-101 could also protect against these complications. Furthermore, understanding CVB immune responses may inform broader vaccine development against other enteroviruses, whose recurring outbreaks pose an emerging threat (41).

Several limitations should be acknowledged. First, our study was conducted in healthy adults, whereas the primary target population for T1D prevention would be genetically at-risk children. Immune responses in pediatric populations may differ due to age-related variations in immune cell compositions and baseline immunity. Trials in children will address this knowledge gap. Second, our sample size was modest, constrained by the size of the parent Phase I trial. Larger trials are needed to confirm our findings and assess relationships to clinical outcomes. Third, we focused on circulating T-cell responses and did not assess tissue-resident memory populations at mucosal sites where CVB-host interactions occur. Future studies should assess mucosal immunity at CVB entry sites. Finally, long-term follow-up beyond 32 weeks will be important to assess durability of vaccine-induced immunity and the need for recall schedules. Tfh responses showed variable persistence at week 32, with some individuals displaying declining frequencies. However, this transient rise in circulating Tfh cells should not be interpreted as loss of vaccine-induced immunity, as memory lymph node-resident rather than circulating Tfh populations may maintain germinal center reactions and Ab production, as documented for other antiviral vaccines (42).

In conclusion, PRV-101 induces a distinctive and favorable immunological profile, characterized by robust CD4⁺ Tfh activation and minimal CD8⁺ T-cell responses that may be diabetogenic. Vaccine-induced Tfh cells correlate with neutralizing Ab production, providing a cellular immunity support for durable humoral immunity. Sex differences, with females exhibiting superior Tfh responses, highlight the importance of sex-stratified analyses in vaccine trials. The successful priming of *de novo* immunity in CVB-seronegative individuals alongside boosting of pre-existing immunity in CVB-seropositive participants support broad applicability. Altogether, these findings encourage continued clinical development of PRV-101 as a safe and immunogenic vaccine candidate for the prevention of CVB-associated diseases, including T1D.

## Data Availability

All data produced in the present study are available upon reasonable request to the authors

## ACKNOWLEDGEMENTS

This study was supported by EU H2023 grant 101137457 (ENT1DEP) (H.H., R.M.), research funding from Provention Bio Inc. (R.M.), PhD fellowship from the *Ministère de l’Enseignement Supérieur et de la Recherche* and *Aide aux Jeunes Diabétiques* (O.B.M.), and a grant from Sigrid Juselius Foundation (H.H.).

## AUTHORS’ RELATIONSHIPS AND ACTIVITIES

H.H. and M.K are board members and stock owner in Vactech Ltd., which develops vaccines against picornaviruses and owns PRV-101 vaccine-related intellectual property rights. F.L. and M.S. were employees of Provention Bio Inc. H.H. has served on the scientific advisory board of Provention Bio Inc. (acquired by Sanofi in 2023). R.M. received research funding for this study from Provention Bio Inc., who had no role in study design, data collection, data analysis, data interpretation, manuscript preparation, or the decision to publish. All other authors declare they have no competing interests.

## CONTRIBUTION STATEMENT

Conceptualization: FV, JEL, HH, RM

Methodology: FV, OBM, RM

Resources: JEL, MSc, MK, FL, MS, HH, SY, RM

Investigation: FV, MP, OBM, JEL, RM

Formal analysis: FV, MP, RM

Visualization: FV, RM

Supervision: FV, HH, SY, RM

Funding acquisition: HH, RM

Project administration: JEL, HH, SY, RM

Writing - original draft: FV, RM

Writing - review & editing: FV, JEL, HH, SY, RM

