## Supplementary material for "PRV-101 Coxsackievirus B vaccine elicits protective T follicular helper immunity while avoiding cytotoxic T-cell responses in humans: implications for type 1 diabetes prevention"

F. Vecchio et al.

### Supplementary figures

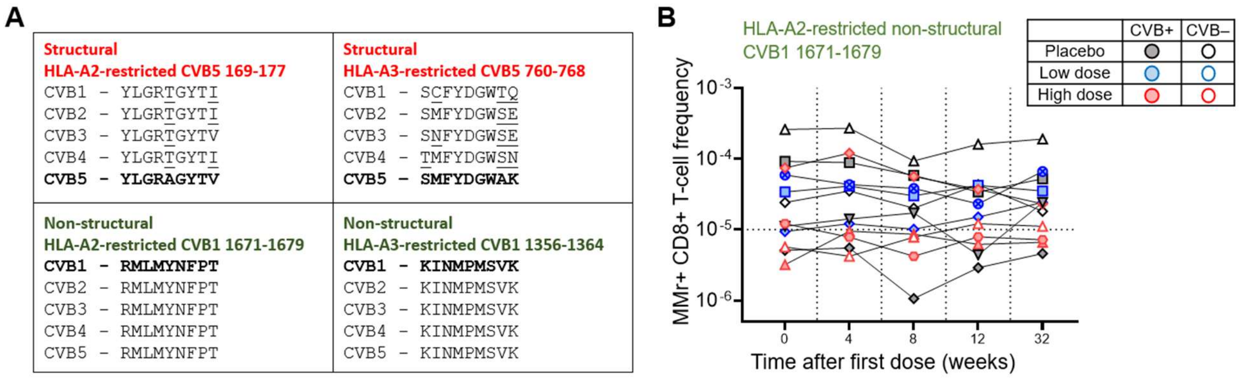


**Supplementary Fig. 1 CVB structural and non-structural epitopes used for CD8+ T-cell analyses and responses to the HLA-A2-restricted non-structural epitope used.** (**A**) Conservation of HLA-A2- and HLA-A3-restricted structural (red) and non-structural epitopes (green) across CVB serotypes 1-5. The reference sequence used is indicated in bold, amino acid differences in other serotypes are underlined. (**B**) Longitudinal frequencies of multimer$^{+}$CD8$^{+}$ T cells recognizing the HLA-A*02:01-restricted non-structural epitope CVB1$_{1671\text{-}1679}$ at weeks 0, 4, 8, 12, and 32. Each symbol represents one participant, with filled and open symbols indicating CVB-seropositive and CVB-seronegative participants at baseline, respectively.

### Supplementary tables

| Participant ID | Treatment arm | Baseline CVB serostatus | Symbol | Sex (M/F) | HLA-A2 | HLA-A3 |
|---|---|---|---|---|---|---|
| 1 | placebo | + | ● | F | | x |
| 2 | placebo | + | ■ | F | x | |
| 3 | placebo | - | ◇ | M | x | |
| 4 | placebo | - | △ | M | x | |
| 5 | placebo | + | ▽ | M | x | |
| 6 | placebo | + | ⊗ | F | | x |
| 7 | placebo | - | ○ | M | | x |
| 8 | placebo | + | ◆ | M | x | |
| 9 | low dose | + | ■ | M | x | |
| 10 | low dose | + | ● | M | | x |
| 11 | low dose | + | ▲ | M | | x |
| 12 | low dose | + | ▼ | M | | x |
| 13 | low dose | + | ◆ | F | x | |
| 14 | low dose | - | ○ | F | x | x |
| 15 | low dose | + | ⊗ | F | x | |
| 16 | high dose | - | ○ | M | | x |
| 17 | high dose | + | ■ | M | | x |
| 18 | high dose | + | ◇ | F | x | |
| 19 | high dose | + | △ | F | x | |
| 20 | high dose | + | ▽ | F | | x |
| 21 | high dose | + | ⊗ | F | | x |
| 22 | high dose | + | ● | M | | x |
| 23 | high dose | - | □ | F | | x |
| 24 | high dose | - | △ | F | x | |
| 25 | high dose | + | ⊠ | F | | x |
| 26 | high dose | + | ⬡ | F | x | |

**Supplementary Table 1. Participants included in the ancillary T-cell study.** Columns display participant ID, treatment arm, baseline CVB serostatus, corresponding symbols used in all figures, sex, HLA-A2 (A*02:01) and HLA-A3 (A*03:01) typing at baseline.
